# The associations of long-term physical activity in adulthood with later biological ageing and all-cause mortality – a prospective twin study

**DOI:** 10.1101/2023.06.02.23290916

**Authors:** Anna Kankaanpää, Asko Tolvanen, Laura Joensuu, Katja Waller, Aino Heikkinen, Jaakko Kaprio, Miina Ollikainen, Elina Sillanpää

## Abstract

**Objectives:** The association between leisure-time physical activity (LTPA) and a lower risk of mortality is susceptible to bias from multiple sources. We investigated the potential of biological ageing to mediate the association between long-term LTPA and mortality and whether the methods used to account for reverse causality affect the interpretation of this association.

**Methods:** Study participants were twins from the older Finnish Twin Cohort (*n*=22,750; 18–50 years at baseline). LTPA was assessed using questionnaires in 1975, 1981 and 1990. The mortality follow-up lasted until 2020 and biological ageing was assessed using epigenetic clocks in a subsample (*n*=1,153) with blood samples taken during the follow-up. Using latent profile analysis, we identified classes with distinct longitudinal LTPA patterns and studied differences in biological ageing between these classes. We employed survival models to examine differences in total, short-term and long-term all-cause mortality, and multilevel models for twin data to control for familial factors.

**Results:** We identified four classes of long-term LTPA: sedentary, moderately active, active and highly active. Although biological ageing was accelerated in sedentary and highly active classes, after adjusting for other lifestyle-related factors, the associations mainly attenuated. Physically active classes had a maximum 7% lower risk of total mortality over the sedentary class, but this association was consistent only in the short term and could largely be accounted for by familial factors. LTPA exhibited less favourable associations when prevalent diseases were exclusion criteria rather than covariate.

**Conclusion:** Being active may reflect a healthy phenotype instead of causally reducing mortality.

## INTRODUCTION

The association between leisure-time physical activity (LTPA) and a lower risk of mortality from all causes and cardiovascular diseases is frequently reported[1–4]. However, the evidence is generally based on observational studies, and LTPA is typically assessed at single time points. Studies using repeatedly measured LTPA have suggested that being both persistently and increasingly active over adulthood are associated with a reduced risk of mortality[2]. However, evidence based on randomized controlled trials has failed to confirm that LTPA prevents premature mortality[5], while genetically informed studies have suggested that genetic selection may partly account for these associations, as genetically healthy participants tend to engage in LTPA[6,7]. In fact, the association between LTPA and mortality is susceptible to reverse-causality bias: an underlying suboptimal physiological or predisease state may negatively affect LTPA, which means that the observed association may be due to a causal relationship between the covert disease state and subsequent premature death[8]. Previous studies have shown that increased control for reverse causality results in weaker associations between LTPA and mortality[8,9]. Moreover, researchers have proposed that, rather than LTPA *per se* reducing mortality risk, participation in LTPA and the ability to increase LTPA in later life are themselves indicators of good fitness and health[10].

Slower biological ageing is a plausible mechanism for explaining the path from an active lifestyle (or a healthy phenotype) to reduced mortality risk. Biological ageing is the gradual and progressive decline in system integrity that occurs with advancing age and results in increased risk of morbidity and mortality[11,12]. Epigenetic clocks produce estimates for biological ageing based on DNA methylation (DNAm) alterations within specific CpG (a cytosine nucleotide followed by a guanine nucleotide) sites and are one of the primary hallmarks of biological ageing[12]. Epigenetic clocks can sum up genetic influences and lifetime burden of lifestyles that predict time-to-death. Although previous studies have suggested that LTPA is associated with decelerated biological ageing[13,14], more research is required because reliable biological age indicators have only recently become available[15–17].

The main purpose of this study was to identify classes of long-term LTPA patterns and to examine whether these classes differ in terms of biological ageing. Furthermore, we aimed to explore class-specific differences in all-cause mortality, considering biological ageing to be a potential mediator of the favourable associations between long-term LTPA and all-cause mortality. To evaluate the influence of reverse causality, we studied whether the association between long-term LTPA and mortality is affected by how prevalent diseases are accounted for, and we investigated the association separately for short-term and long-term mortality. Finally, using a twin study design, we examined whether the associations between long-term LTPA and mortality are independent of shared genetic and environmental factors.

## MATERIAL AND METHODS

### Study design and participants

Participants were twins from the older Finnish Twin Cohort (FTC)[18]. The FTC consists of same-sex twins born in Finland before 1958 and with both co-twins alive in 1967. Questionnaires were mailed in 1975 and 1981 to all twins born before 1958 and living in Finland, with a follow-up questionnaire conducted in 1990 with twins born in 1930–1957. The response rates were high (77–89%). Participants aged 18–50 years at baseline in 1975, who had at least one measurement of LTPA and were alive in 1990, were included in the present study (*n*=22,750). For the subsample (*n*=1,153), blood samples were taken during the 1993–2020 period, and blood-based DNAm was used to assess biological ageing at ages ranging from 37 to 81 years.

### Measurements

#### Leisure-time physical activity (LTPA)

LTPA was measured in metabolic equivalent (MET) hours per day (h/day) using a structured validated questionnaire in 1975, 1981 and 1990[7,19–21] (see online supplementary material and Tables S1–S2 for details).

#### Outcome variables

*Biological ageing* was assessed using blood-based epigenetic ageing measures, namely, principal component (PC)–based DNAm GrimAge[16,17] and DunedinPACE[15]. The measurement and preprocessing of the DNAm levels, as well as additional information on the epigenetic ageing measures, are provided in the online supplementary material. To summarise, DNAm GrimAge is a mortality predictor by design, and it is composite of age, sex, DNAm-based surrogates for seven plasma proteins and smoking pack-years[17]. Recently, PC-based epigenetic clocks have been developed to bolster the reliability and validity of the clocks[16]. We produced PC-based GrimAge estimates using an R package ([16], https://github.com/MorganLevineLab/PC-Clocks). Age acceleration (AA_PC-Grim_) in years was determined as a residual by regressing the estimated epigenetic age on chronological age. In addition, we obtained PC-based components of GrimAge (adjusted for age), including DNAm smoking pack-years, DNAm adrenomedullin (ADM), DNAm beta-2-microglobulin (B2M), DNAm cystatin C, DNAm growth differentiation factor 15 (GDF15), DNAm leptin, DNAm plasminogen activator inhibitor 1 (PAI-1), and DNAm tissue inhibitor metalloproteinases 1 (TIMP-1).

The DunedinPACE estimator provides an estimate of the pace of ageing in years per calendar year; the estimates were calculated using an R package ([15], https://github.com/danbelsky/DunedinPACE).

##### Mortality

Dates of death were retrieved from the Population Register Centre of Finland and Statistics Finland. The mortality follow-up started with the response date to the 1990 questionnaire and continued until the date of emigration, death or 31 December 2020, whichever came first.

#### Confounding variables

Specific diseases and other lifestyle-related factors that could have affected long-term LTPA and mortality were considered as potential confounders (see online supplementary material).

##### Health status

An indicator of specific physician-diagnosed somatic diseases (angina pectoris, myocardial infarction, and diabetes) based on self-reports from 1975 and 1981 was used.

*Education* (in years) was based on the self-reported latest education in 1981 and converted into years of education as follows: less than primary school (3), primary school (6), junior high school (9), high school graduate (12), university degree (16), and ≥ 1 year of education such as vocational training in addition to primary school (7), junior high school (10) or high school (13)[22].

*Body mass index (BMI)* (kg/m^2^) was calculated based on self-reported height and weight in 1981. BMI based on self-reports has been shown to agree well with BMI based on measured values[23].

*Smoking status* was self-reported based on an extensive smoking history[24] and classified as never, occasional, former and current light (1–9 cigarettes per day [CPD]), medium (10–19 CPD) and heavy (≥20 CPD) smokers.

*Alcohol use* was based on average alcohol consumption (g/day) in 1981 of beer, wine and spirits[25] and classified as never, former, occasional (>0.1 and <1.3 g), low (≥1.3 and <25 g), medium (≥25 and <45 g), high (≥45 and <65 g) and very high (≥65 g)[26].

### Statistical analysis

The main analyses were conducted using Mplus 8.2[27]. The parameters of the models were estimated using the full information maximum method (FIML) with robust standard errors, which uses all available data during estimation.

We conducted latent profile analysis (LPA) to identify long-term patterns of LTPA based on the means and variances of MET indices in 1975, 1981 and 1990. An LPA model with 1–6 classes was fitted (see online supplementary material). The mean differences between the classes in later biological ageing were studied using the Bolck-Croon-Hagenaars (BCH) approach, which controls for measurement error in classification[28]. The standard errors of the models were corrected for nested sampling within families. The analyses were adjusted for sex, age and health status (Model 1) as well as for education, BMI, smoking and alcohol use (Model 2). In addition, differences in the DNAm-based plasma proteins and smoking pack-years between the classes were explored.

We investigated differences in total all-cause mortality as well as in short- and long-term all-cause mortality between the classes using a discrete-time survival model, which enables flexible modelling within the structural equation framework[29,30] (see online supplementary material and Figure S1). Although the model was adjusted using a procedure similar to that employed for the models of biological ageing, we accounted for health status in two ways. First, we adjusted the model for health status, and second, we excluded participants who reported selected diseases (angina pectoris, myocardial infarction, and diabetes) from the analysis.

To account for familial factors, the associations between long-term LTPA and total all-cause mortality were studied at the within-twin-pair level using multilevel modelling. For monozygotic (MZ) twin pairs, the association was controlled for shared environmental factors and genetics, while for dizygotic (DZ) pairs, it was controlled for shared environmental factors and partially for genetics.

## RESULTS

The descriptive statistics of the study variables are presented in Table 1 and are stratified by sex in the online supplementary Table S3.

**Table 1.**
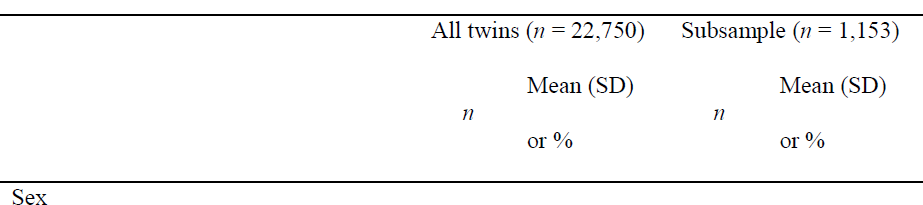
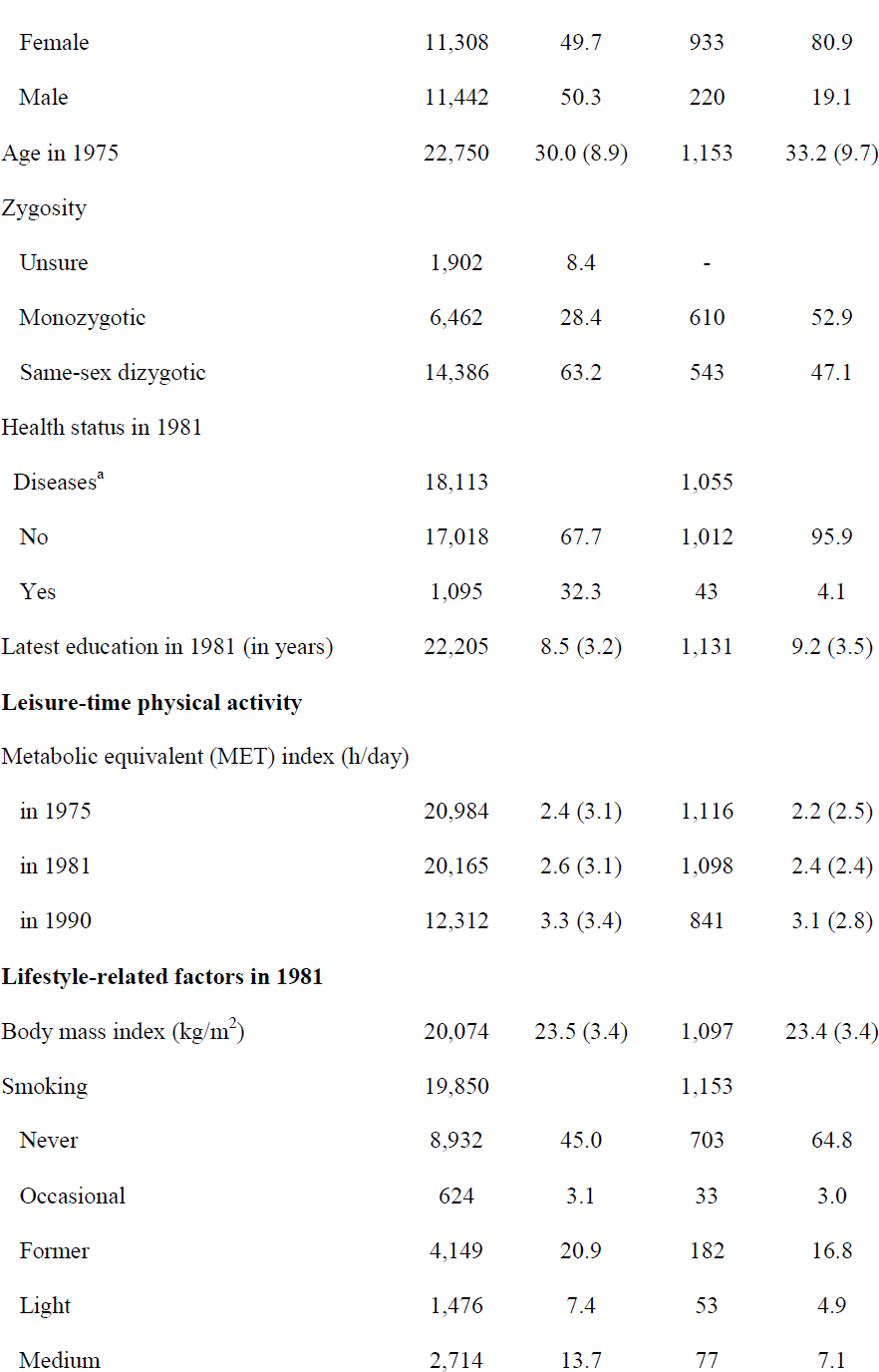
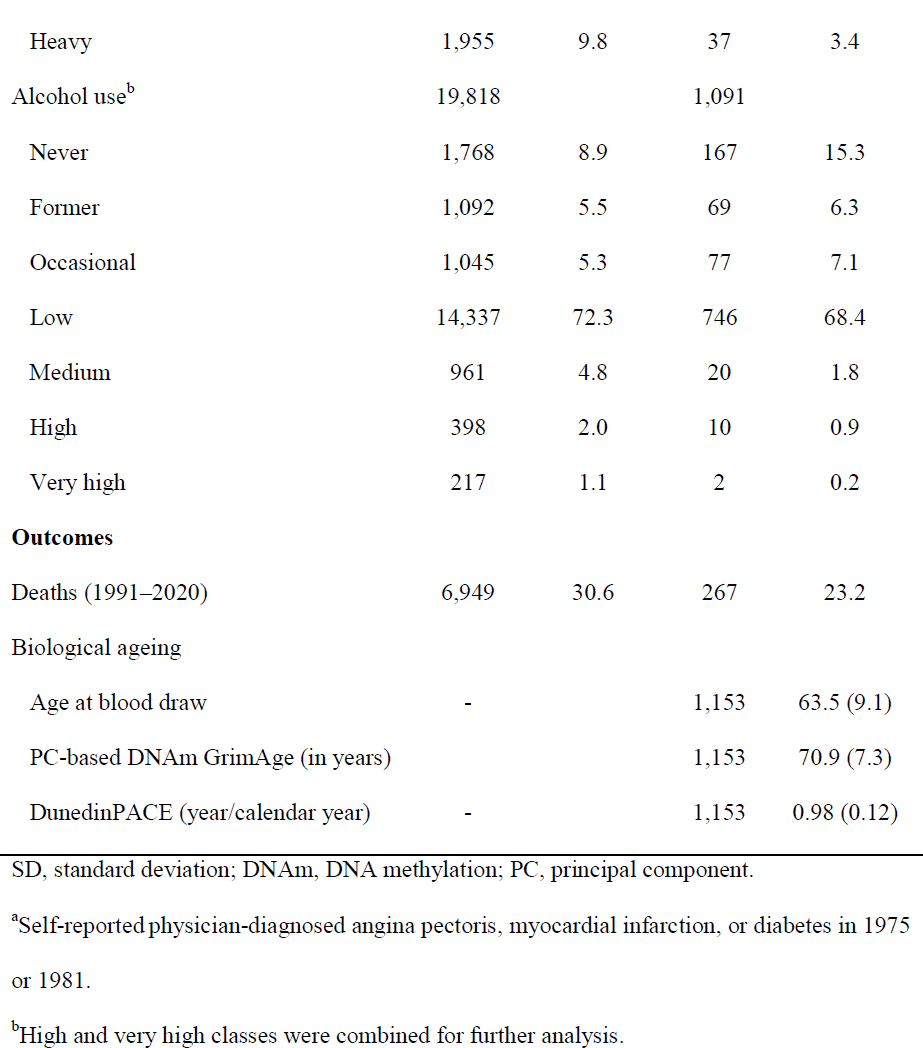
Descriptive statistics of the study variables for all twins and the subsample of twins with information on biological ageing

### Patterns of long-term LTPA

We identified four classes with different long-term LTPA patterns (Figure 1). The model selection for and characteristics of the long-term LTPA classes are described in online supplementary material and Tables S4–S5. For sensitivity analysis, we conducted the main analyses using a five-class solution (online supplementary material and Figures S4–S6).

**Figure 1.**
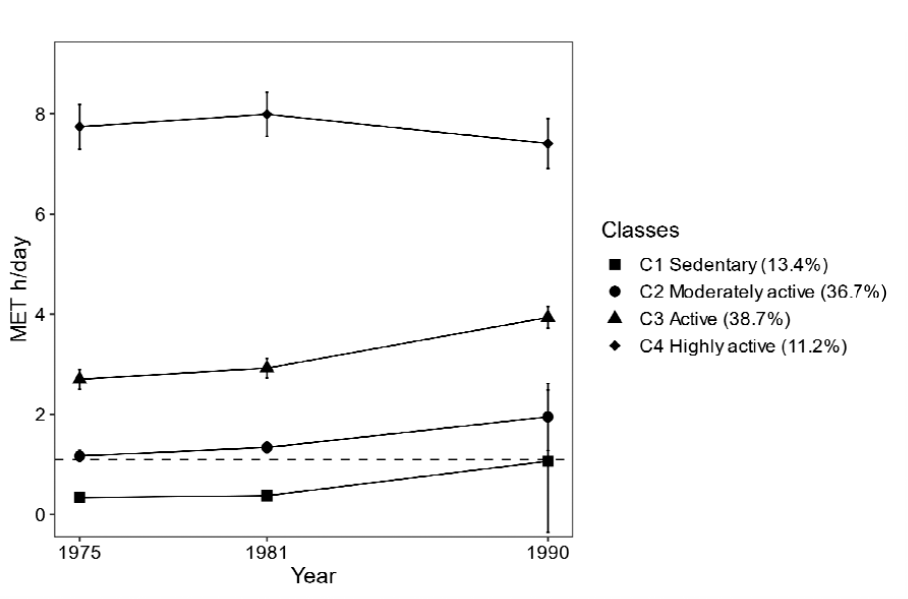
Patterns of long-term leisure-time physical activity (*n* = 22,750). Means of metabolic equivalent (MET) hours (h)/day and 95% confidence intervals are presented. The dashed line denotes World Health Organization guidelines for the recommended minimum amount of physical activity for adults (150 min of moderate intensity physical activity per week ∼ 1.1 MET h/day)[31].

### Differences in biological ageing between long-term LTPA classes

There were differences between the classes in terms of biological ageing, measured using AA_PC-Grim_ and DunedinPACE (Figure 2). The association between long-term LTPA and biological ageing followed a U-shaped pattern: participants in the sedentary and highly active classes were biologically older than those who were moderately active and active. After adjusting for other lifestyle-related factors, most differences were attenuated, but based on AA_PC-Grim_, the highly active class remained, on average, 1.3 years (95% confidence interval: 0.3–2.3) biologically older than the moderately active class and 1.8 years (0.7–2.8) biologically older than the active class. As no significant beneficial association between long-term LTPA and slower biological ageing was observed, biological ageing was unlikely to act as a mediator for the association between long-term LTPA and lower mortality, and no further path modelling was conducted. Of the PC-GrimAge components, the U-shaped association was most pronounced in DNAm-based cystatin C and B2M (see online supplementary material and Figure S2).

**Figure 2.**
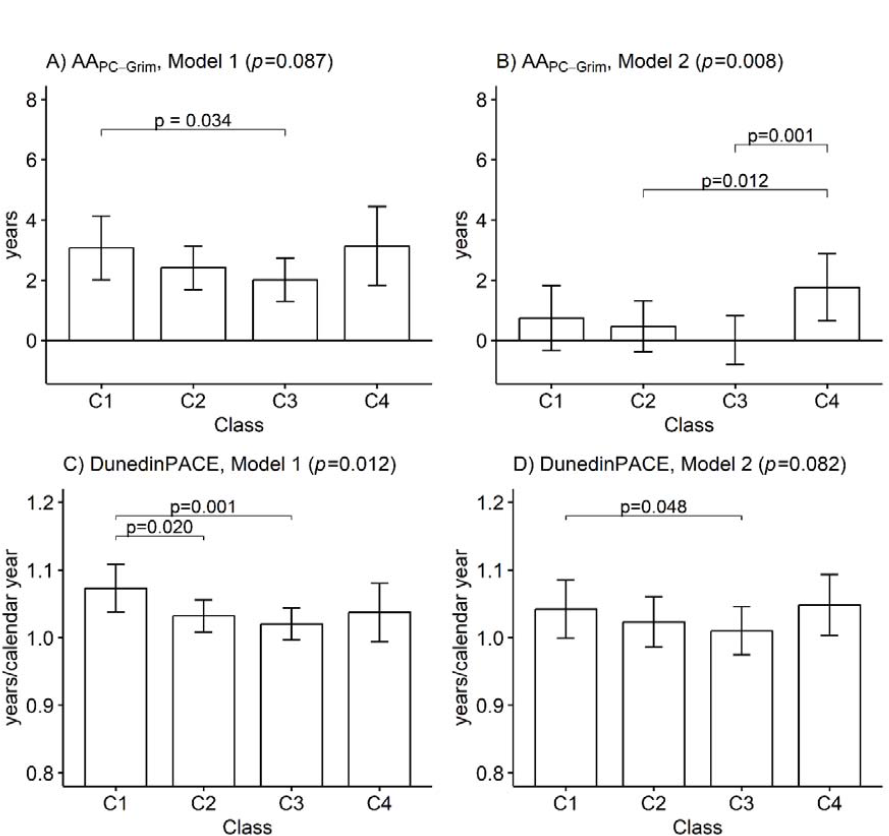
Mean differences between the long-term leisure-time physical activity classes in terms of biological ageing measured using A)–B) PC-based GrimAge and C)–D) DunedinPACE (*n* = 1,153). Means and 95% confidence intervals are presented. Model 1 was adjusted for sex, age and health status, and Model 2 was additionally adjusted for education, body mass index, smoking and alcohol use. C1: Sedentary (8.8%); C2: Moderately active (38.4%); C3: Active (45.5%); C4: Highly active (7.3%); AA, Age acceleration; *p*-values from the Wald test.

### Differences in mortality between the latent long-term LTPA classes

Over a third (38.8%) of the participants from the sedentary class died during the mortality follow-up period, compared to 30.8%, 29.0% and 25.4% from the more active classes, respectively. Active classes had 15–23% lower all-cause mortality risk compared to the sedentary class, but after accounting for other health-related factors, the reduction in mortality risk was a maximum of 7% (Figure 3A).

**Figure 3.**
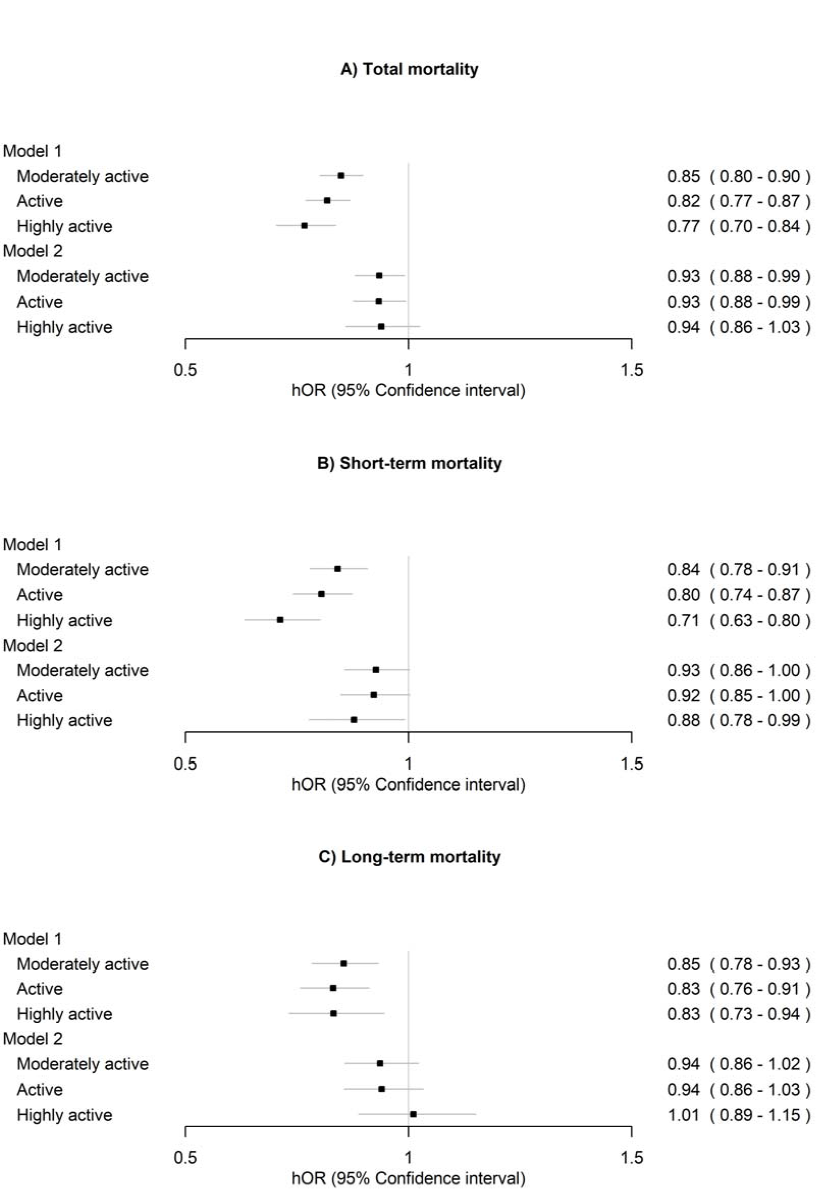
Associations of long-term leisure-time physical activity with A) total mortality, B) short-term mortality (1990–2011) and C) long-term mortality (2012–2020) (*n* = 22,750). The sedentary class was treated as the reference. Model 1 was adjusted for sex (female), age and health status. Model 2 was additionally adjusted for education, body mass index, smoking and alcohol use. hOR, hazard odds ratio.

Approximately half of the deaths occurred in 2011 or earlier; 2011 was considered the cut-off point for short- and long-term mortality (Figure 3B–3C). For sensitivity analysis, we performed the modelling using 2006 as the cut-off, when one-third of the deaths had occurred (online supplementary Figure S7).

Overall, the favourable associations of long-term LTPA were more consistent with short-term than long-term mortality. In particular, high activity was associated only with lower short-term mortality. The results were similar after excluding participants with reported diseases (online supplementary Figure S3).

### Within-twin-pair differences in mortality between long-term LTPA classes

In a model adjusted only for health status at the within-pair level, the analyses showed that the moderately active and active classes exhibited lower risks of all-cause mortality compared to the sedentary class within all pairs, DZ pairs and MZ pairs (Figure 4, Model 1). After additionally adjusting the model for other lifestyle-related factors, the differences were considerably attenuated, but the moderately active and active classes exhibited lower risks of all-cause mortality compared to the sedentary class within all pairs and MZ pairs (Model 2).

**Figure 4.**
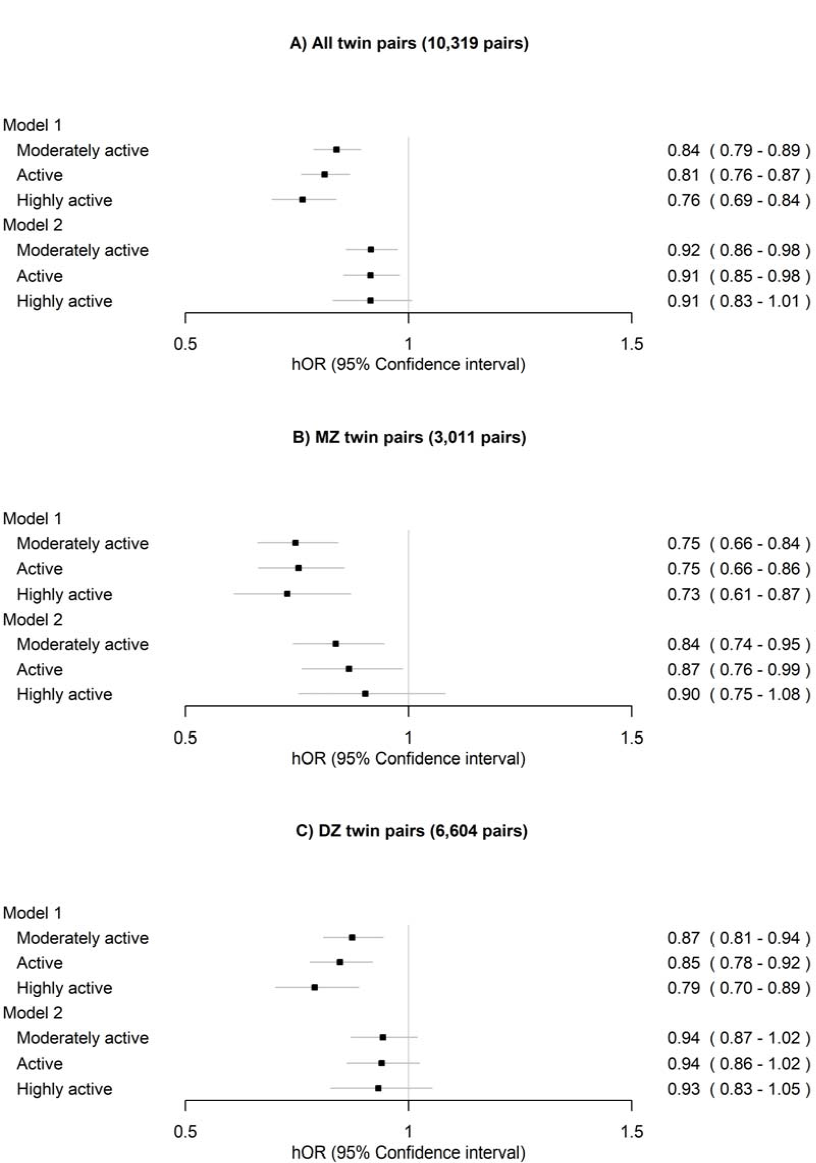
Within-twin-pair differences in all-cause mortality between the long-term leisure-time physical activity classes for A) all twin pairs, B) monozygotic (MZ) pairs and C) dizygotic (DZ) pairs. The sedentary class was treated as the reference. Only twin pairs with information on LTPA and alive in 1990 were included in the analysis. Model 1 was adjusted for sex (female) and age at the between-twin-pair level and health status at the within-twin pair level. Model 2 was additionally adjusted for education, body mass index, smoking and alcohol use at the within-twin pair level. hOR, hazard odds ratio.

After excluding the twin pairs who reported specific diseases, the differences in all-cause mortality were almost fully attenuated and were no longer significant after adjusting for other lifestyle-related factors for all twin pairs, MZ pairs and DZ pairs (Figure 5, Model 2).

**Figure 5.**
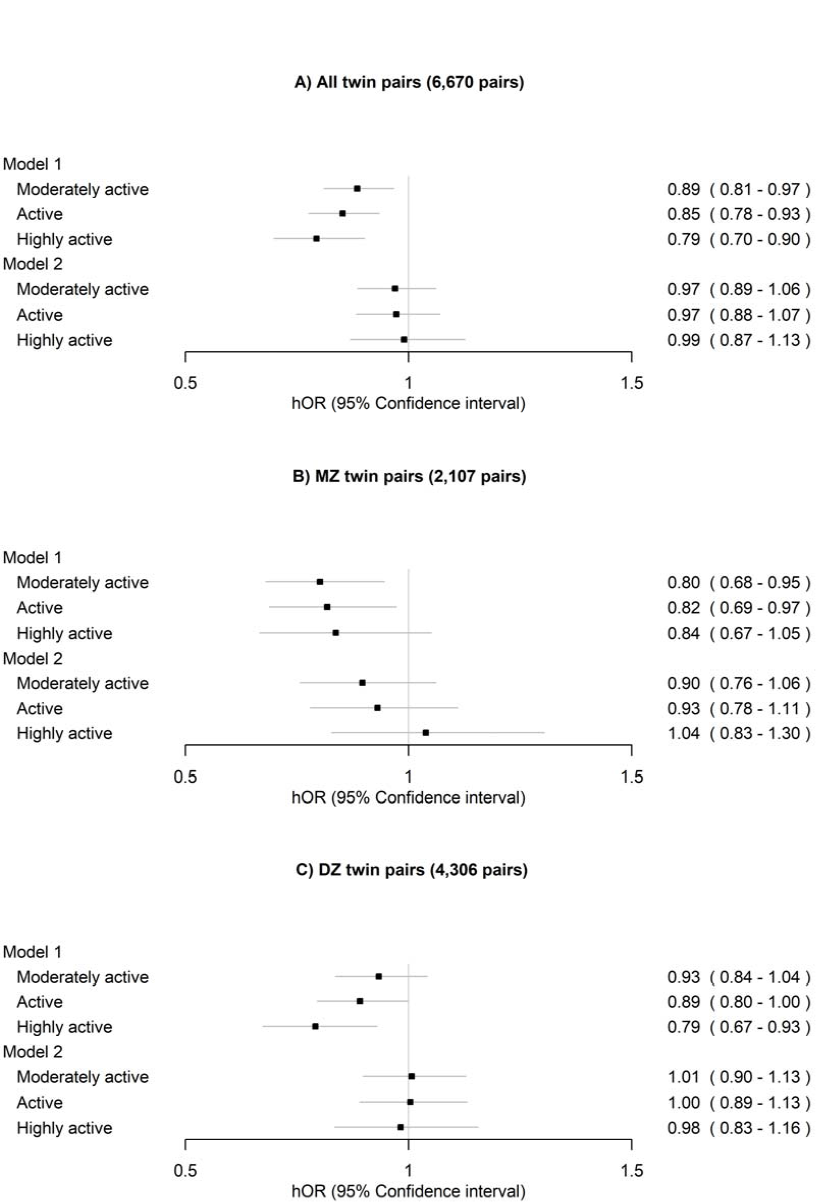
Within-twin-pair differences in all-cause mortality between the long-term leisure-time physical activity classes for A) all twin pairs, B) monozygotic (MZ) pairs and C) dizygotic (DZ) pairs after excluding twin pairs who reported specific diseases. The sedentary class was treated as the reference. Only twin pairs with information on LTPA and alive in 1990 were included in the analysis. Model 1 was adjusted for sex (female) and age at the between-twin-pair level. Model 2 was additionally adjusted for education, body mass index, smoking and alcohol use at the within-twin-pair level.

## DISCUSSION

We conducted a long-term mortality follow-up of a longitudinal study of LTPA with a large cohort of adult twins and identified classes according to long-term LTPA patterns. Our analyses were based on standard cohort approaches to individuals (adjusting for the sampling of twin pairs) and on within-pair modelling to account for familial and genetic factors. The results showed that the beneficial associations of long-term LTPA with slow biological ageing and reduced mortality were largely accounted for by other health-related factors. The most remarkable reduction of 7% in all-cause mortality was observed already when the recommended minimum amount of LTPA was, on average, achieved, with no additional benefits provided by higher levels of LTPA. This result is in line with the World Health Organization guidelines[31] and studies demonstrating that dose-response association between physical activity (PA) and all-cause mortality is curvilinear rather than linear[3,4]. The association between long-term LTPA and all-cause mortality was susceptible to reverse-causality bias because a consistent association was observed only in the short term. Moreover, the results of within-twin-pair comparisons depended on how health status was controlled for. An alternative explanation is a change in PA exposure as a function of time; in other words, some people may have changed their PA levels later in adulthood.

### The association between long-term LTPA and mortality was susceptible to bias from multiple sources

Previous studies of long-term LTPA have reported larger differences in mortality between consistently active and inactive participants (16–36%)[32–34] than what we observed in our study. This discrepancy can be explained by differences in the length of the mortality follow-up time, which has been shown to affect associations between PA and mortality[8]. In our study, participants were followed over a 30-year period; therefore, we were able to divide the follow-up time into two parts to reflect short- and long-term survival. Our analysis revealed that the associations of long-term LTPA were more consistent with short-term than long-term mortality. Being highly active was associated with reduced mortality only in the short term and thus may not have long-term mortality benefits unless activity is maintained continuously.

Another reason for the higher effect sizes observed in previous studies may be residual confounding due to insufficient adjustments. In our study, when the models were minimally adjusted, the reduction in mortality (15–23%) was closer to the level observed in previous studies. After careful adjustment for smoking (in terms of both status and quantity) and other lifestyle-related factors (education, BMI and alcohol use), the association was considerably attenuated. Smoking is the most harmful lifestyle habit in terms of mortality[35,36]. Generally, only smoking status is adjusted for in analyses, but this may not be sufficient because current smokers who are physically active tend to smoke less than those who are sedentary (online supplementary Table S5).

Previous twin studies have provided somewhat inconsistent results regarding the association between LTPA and mortality after accounting for familial factors. Studies using data from Finnish twins have suggested that the association between LTPA and all-cause mortality is due to genetic selection, as there was no difference in mortality between MZ co-twins discordant in terms of LTPA[7,36], whereas a study with a similar twin study setting found a difference within Swedish twin pairs[37]. Several reasons for this discrepancy have been proposed, such as differences in LTPA measurement methods and in controlling for prevalent diseases[38]. Our within-twin-pair comparisons were considerably affected by how the prevalent diseases were controlled for. Excluding twin pairs with one or both co-twins reporting diseases (as done in prior Finnish studies) attenuated the differences more strongly compared to adjusting for health status, particularly when other lifestyle-related factors were controlled for. This finding may support studies arguing that adjusting the survival model for health status may not be sufficient to mitigate reverse causation[9] and may also reflect an accumulation of unhealthy lifestyle habits and prevalent diseases in the sedentary class (online supplementary Table S5).

### Long-term LTPA and biological ageing

Previous studies, mostly based on cross-sectional data, have indicated that the observed associations, or lack of associations, between LTPA and biological ageing depend on the type of epigenetic clock used[13,39–42]. This is probably because first-generation clocks were developed to predict chronological age[43,44], while newer clocks, such as DNAm GrimAge, are better predictors of health and lifespan[17,45]. LTPA is most consistently associated with biological ageing, assessed using the DNAm GrimAge estimator. To the best of our knowledge, this is the first study to report on the association between LTPA and DunedinPACE. We found that biological ageing was, on average, slower in the active class compared to the sedentary class when these two markers of biological ageing were used. However, after adjusting the model for other lifestyle-related factors, the differences were largely attenuated, which likely reflects an accumulation of factors related to an unhealthy lifestyle in the sedentary class[46]. Moreover, the weak associations observed in the present study may be due to the prospective study design, as the beneficial influences of LTPA may diminish after a time lag of several years.

Contrary to the existing literature [13,40,42], we observed that the highly active class was, on average, biologically older than the moderately active and active classes when the DNAm (PC) GrimAge was used. However, a recent study indicated a curvilinear association between accelerometer-based PA and biological ageing[41]. Some studies have reported U-shaped associations between LTPA and mortality[47], and it has been suggested that sudden cardiac deaths after/during exercise may explain the increased mortality at high levels of LTPA[10,47]. However, a recent meta-analysis did not find evidence of increased levels of mortality at high PA levels[1].

To better understand the reasons for the observed patterns, we explored the DNAm-based plasma proteins included in the DNAm GrimAge estimator. Interestingly, the levels of DNAm-based cystatin C and beta-2-microglobulin (B2M) were higher in both the sedentary and highly active classes. These proteins are markers of kidney function, and their higher concentrations are linked to mortality due to cardiovascular diseases[48] and sudden cardiac death[49]. Further studies are required to determine whether these DNAm-based proteins play a role in sudden cardiac death in athletes.

### Strengths and limitations

Our study has major strengths. We used prospective population-based large cohort data with longitudinal measurements of LTPA over 15 years with validated questionnaires and 30 years of mortality follow-up. The LTPA patterns were studied using a data-driven method without using preselected cut-offs. Biological ageing was assessed using novel epigenetic clocks, which have been shown to perform better than their predecessors[15,16]. The twin study design enabled us to control the analyses for genetics and the shared environment.

However, our study also had limitations. LTPA was self-reported, and a considerable proportion of the participants did not have LTPA at all three measurement points. Self-reports may be susceptible to recall and social-desirability biases. However, the questionnaire-based MET index has been shown to be a reliable tool for measuring LTPA[20], and self-reports may better reflect long-term LTPA than device-based measures.

## Conclusions

Our findings support the suggestion that, rather than LTPA per se reducing the risk of mortality, being active may be an indicator of a healthy phenotype and an overall healthy lifestyle, which co-occur with a lower mortality risk. Further research on DNAm-based surrogates may provide insights into the mechanisms behind the beneficial and detrimental health influences of LTPA.

## Contributorship

AK, LJ, KW, MO, JK and ES conceptualised the research question. AH preprocessed the DNAm data. AK, AT and ES designed the statistical analysis, and AK performed the statistical modelling under the supervision of AT. JK and MO designed and collected the FTC dataset and participated in designing the analysis of this manuscript. AK drafted the first version of the manuscript, and ES and LJ contributed significantly to the writing. AK, AT, LJ, KW, AH, JK, MO and ES contributed to the interpretation of the results and revising the manuscript. ES, MO and JK acquired the funding for the study.

## Data sharing

A subsample of the FTC with DNA methylation age estimates, phenotypes and information on the classes of long-term LTPA will be located in the Biobank of the National Institute for Health and Welfare. All these data will be publicly available for use by qualified researchers following a standardised application procedure (for details on the application process, see the following website: https://thl.fi/en/web/thl-biobank/for-researchers). Because of the consent given by the study participants and the high degree of identifiability of the twin siblings in Finland, the full cohort data cannot be made publicly available. However, the full cohort data are available through the Institute for Molecular Medicine Finland (FIMM) Data Access Committee (DAC) for authorised researchers with an IRB/ethics approval and an institutionally approved study plan. For more details, please contact the FIMM DAC.

## Ethical approval

Data collection was conducted in accordance with the Declaration of Helsinki. The ethics committees of the University of Helsinki and Helsinki University Central Hospital approved the study protocol (113/E3/2001 and 346/E0/05). Blood samples for DNA analyses were collected during in-person clinical studies after written informed consent was signed.

## Data Availability

A subsample of the FTC with DNA methylation age estimates, phenotypes and information on the classes of long-term LTPA will be located in the Biobank of the National Institute for Health and Welfare. All these data will be publicly available for use by qualified researchers following a standardised application procedure (for details on the application process, see the following website: https://thl.fi/en/web/thl-biobank/for-researchers). Because of the consent given by the study participants and the high degree of identifiability of the twin siblings in Finland, the full cohort data cannot be made publicly available. However, the full cohort data are available through the Institute for Molecular Medicine Finland (FIMM) Data Access Committee (DAC) for authorised researchers with an IRB/ethics approval and an institutionally approved study plan. For more details, please contact the FIMM DAC (fimm-dac@ helsinki.fi).

