## Supplementary material for "The associations of long-term physical activity in adulthood with later biological ageing and all-cause mortality – a prospective twin study"

**METHODS**

**Measurements**

Leisure-time physical activity (LTPA)

In 1975 and 1981 questions concerned monthly frequency of LTPA, mean duration and intensity of the sessions (Table S1). The MET index was calculated by multiplying the frequency, intensity, and duration of leisure activities as well as commuting activities, and then summing up the resulting values[1,2]. In 1990 the questionnaire slightly differed and participants reported their time spent in LTPA (including commuting activity) at different intensity levels (Table S2). The MET index was calculated by multiplying the time spent in LTPA by the estimated MET value of each intensity level and then summing up the resulting values[3]. The MET index was expressed in MET hours/day.

Outcome variables

*Biological aging*. Genomic DNA was extracted from peripheral blood samples using commercial kits. High molecular weight DNA samples (1 μg) were bisulfite converted using EZ-96 DNA /methylation-Gold Kit (Zymo Research, Irvine, CA, USA) according to the manufacturer's protocol. The twins and co-twins were randomly distributed across plates, with both twins from a pair on the same plate. DNA methylation (DNAm) profiles were obtained using Illumina’s Infinium HumanMethylation450 BeadChip or the Infinium MethylationEPIC BeadChip (Illumina, San Diego, CA, USA). The Illumina BeadChips measure single-CpG resolution DNAm levels across the human genome. With these assays, it is possible to interrogate over 450,000 (450k) or 850,000 (EPIC) methylation sites quantitatively across the genome at single-nucleotide resolution. Of the samples included in the present study, 419 were assayed using 450k and 734 samples using EPIC arrays. Methylation data from different platforms was separately preprocessed using R package *meffil*[4] We calculated detection p-values comparing total signal for each probe to the background signal level, to evaluate quality of the samples[5]. Samples of poor quality (mean detection p > .01) were excluded from further analysis. Data were normalized by using the single-sample Noob normalization method, which is suitable for datasets originating from different platforms[6]. We also used Beta-mixture quantile (BMIQ) normalization[7]. Beta values representing CpG methylation levels were calculated as ratio of methylated intensities (M) to the overall intensities (Beta value=M/(M+U+100), where U is unmethylated probe intensity). These preprocessed Beta values were used as input in the calculations of the estimates of epigenetic aging.

DNAm GrimAge is mortality predictor by design[8]. DNAm GrimAge, includes 1 030 CpG sites and was a product of the 2-step development method. It first utilized DNAm data to predict a set of biomarkers (plasma proteins and smoking pack-year) and then these developed DNAm-based biomarkers were used to predict all-cause mortality. In both steps, information on participants’ sex and chronological age was used as well.

DunedinPACE estimator gives an estimate for pace of aging in years per calendar year[9]. DunedinPACE was trained to predict composite measure of pace of aging which describes longitudinal changes over time in several biomarkers of organ-system integrity among same-aged individuals. Pace of aging includes changes in 19 biomarkers measured at four time points over ages from 26 to 45.

Initially, the original DNAm GrimAge estimator was used to produce epigenetic age estimates in years were utilizing calculator ([https://dnamage.genetics.ucla.edu/new)](https://dnamage.genetics.ucla.edu/new) and the age acceleration (AA_Grim_) was defined as the residual from regressing the epigenetic age on chronological age. The measures were screened for outliers (> 5 standard deviations away from mean). There were 16 outliers when using DNAm GrimAge, and none when using DunedinPACE and PC-based measure (AA_PC-Grim_) were used. Therefore PC-based measure was chosen instead of the original DNAm GrimAge estimator for the further analyses.

Confounding variables

Time-varying confounders cause bias for the studies on long-term LTPA and mortality[10]. In our study, on one hand, LTPA was measured over 15-year period and the other lifestyle-related factors may have considerably changed after baseline. On the other hand, exposure may have affected the other lifestyle-related factors measured over a long period and thus, these factors may partly mediate rather than confound the association between exposure and outcome[10]. Moreover, our sample included older participants (born in 1925–29) who had information on mortality (n=1,667) and DNAm (n=144) but were not invited to answer questionnaire in 1990. For these reasons, we considered year 1981 the optimal measurement point of confounders.

**Statistical analysis**

Patterns of long-term LTPA

Several indices were used to evaluate the goodness of fit: Akaike’s information criterion, Bayesian information criterion and sample size-adjusted Bayesian information criterion. The lower values of the information criteria indicated a better fit for the model. Furthermore, we used the Vuong–Lo–Mendell–Rubin likelihood ratio (VLMR) test and the Lo–Mendell–Rubin (LMR) test to determine the optimal number of classes. The estimated model was compared with the model with one class less, and the low *p* value suggested that the model with one class less should be rejected. At each step, the classification quality was assessed using the average posterior probabilities for most likely latent class membership (AvePP). AvePP values close to 1 indicate a clear classification. In addition to the model fit, the final model for further analyses was chosen based on the parsimony and interpretability of the classes.

Discrete-time survival models

Because BCH approach was not available for the survival models, we used posterior classification probabilities as sampling weights to control for measurement error in the classification. We used year as the unit of discrete-time survival indicators from 1991 to 2020 and constructed a latent variable describing propensity for death[11,12] (online Supplementary figure S1A). The latent variable was regressed on the latent class membership of long-term LTPA and the potential confounding variables. Moreover, we divided the follow-up time into two parts, formed latent variables representing propensity for short- and long-term death, and the associations of long-term LTPA on short- and long-term mortality were studied (online Supplementary figure S1B). This approach relaxes strict proportionality assumption often necessary in the survival modeling, by allowing the associations of exposure with short- and long-time mortality to vary. The standard errors of the model parameters were corrected for nested sampling within families.

**RESULTS**

**Patterns of long-term LTPA**

The model-fit based on the information criteria improved at each step (online supplementary Table S4). However, at the sixth step, only a small class (<5%) was extracted, and therefore, a solution with 4–5 classes was considered optimal. At the fifth step, a class with increasing LTPA pattern from sedentary to moderate level was identified. To achieve sufficient power for the further analysis, we used a four-class solution in the main analyses (Figure 1). The level of LTPA appeared to increase slightly between years 1981 and 1990, except in the highly active class. This is probably due to the small changes in the questionnaire (see online supplemental tables S1–2) rather than reflecting actual increase in LTPA.

**Differences in biological aging between the classes of long-term LTPA**

Differences in DNAm-based plasma proteins and smoking pack-years

The overall test for differences between the classes of long-term LTPA in DNAm based plasma proteins indicated differences for DNAm B2M and cystatinC (Figure S2). The mean profiles of these proteins appeared to follow U-shaped pattern; The highest levels were observed in the sedentary and highly active classes. There were also differences in DNAm-based smoking pack-years (Figure S2). In the highly active class, the level was higher than in the other classes, which may indicate under-reporting in the highly active class because the models were adjusted for smoking. There were no differences in DNAm ADM, GDF15, leptin, PAI-1 or TIMP-1.

**Sensitivity Analysis using a five-class solution**

After including fifth class in the LCA model, a class of increasingly active (from sedentary to moderate) participants was extracted (Figure S4). According to the overall test there were differences between the classes in biological aging measured with AA_PC-Grim_ but not in DunedinPACE (Figure S5). Biological aging measured with AA_PC-Grim_ appeared to be accelerated in highly active class. There were differences in mortality between the classes, but the differences were smaller than observed in the main analysis (Figures S6). Increasingly active class did not differ from sedentary class in terms of mortality risk. After accounting for other health-related factors, there were differences only in short-term mortality.

| Table S1. Physical activity questionnaire in 1975 and 1980 and scoring for MET index. | |
| --- | --- |
| **LEISURE ACTIVITY** |  |
| **Intensity** |  |
| Is your physical activity during leisure-time about as strenuous on average as: | |
|  | Score (MET) |
| a) walking | 4 |
| b) alternately walking and jogging | 6 |
| c) jogging | 10 |
| d) running | 13 |
| **Duration** |  |
| How long does the physical activity last at one session on average? | |
|  | Score (min) |
| a) Less than 15 minutes | 7.5 |
| b) 15 min – less than 30 min | 22.5 |
| c) 30 min – less than 1 hour | 45 |
| d) 1 hour – less than 2 hours | 90 |
| e) Over two hours | 120 |
| **Frequency** |  |
| Presently how many times per month do you engage in physical activity during your leisure time? | |
|  | Score (times per month) |
| a) less than once a month | 0.5 |
| b) 1-2 times per month | 1.5 |
| c) 3-5 times per month | 4 |
| d) 6-10 times per month | 8 |
| e)11-19 times per month | 15 |
| f) more than 20 times per month | 20 |
| **COMMUTING ACTIVITY** |  |
| **Intensity** |  |
|  | Score (MET) |
|  | 4 |
| **Duration** |  |
| How much of your daily journey to work is spent in walking, cycling, running and/or cross-country skiing? | |
|  | Score (min) |
| a) Less than 15 minutes | 7 |
| b) 15 min – less than a half an hour | 22 |
| c) half an hour to less than an hour | 45 |
| d) hour or more | 75 |
| e) I am presently not at work | 0 |
| **Frequency** |  |
|  | Score (times per month) |
|  | 20 (5×4) |

| Table S2. Physical activity questionnaire in 1990 and scoring for metabolic equivalent (MET) index. | |
| --- | --- |
| Following question are about your physical activity during leisure time or during your daily journey to work during last 12 months. How many hours in week you engage in physical activity corresponding to each intensity level? | |
| **Intensity levels** | Score (MET) |
| Walking | 4 |
| Alternately walking and jogging | 6 |
| Jogging | 10 |
| Running | 13 |
| **Duration** |  |
|  | Score (min per month) |
| a) Not at all | 0 |
| b) Less than 30 min per week | 60 (15×4) |
| c) 30 min – less than 1 hour per week | 180 (45×4) |
| d) 2-3 hours per week | 600 (150×4) |
| e) 4 hours or more per week | 960 (240×4) |

| Table S3. Descriptive statistics of the study variables stratified by sex for all twins and the subsample of twins with information on biological aging. | | | | | | | | |
| --- | --- | --- | --- | --- | --- | --- | --- | --- |
|  | All twins (n = 22,750) | |  |  | Subsample (n = 1,153) | |  |  |
|  | Men (n = 11,308) | | Women (n = 11,442) | | Men (n = 220) | | Women (n = 933) | |
|  | n | Mean (SD) or % | n | Mean (SD) or % | n | Mean (SD) or % | n | Mean (SD) or % |
| Age in 1975 | 11,308 | 30.2 (8.9) | 11,442 | 29.8 (9.0) | 220 | 26.2 (5.6) | 933 | 34.8 (9.7) |
| Zygosity |  |  |  |  |  |  |  |  |
| Unsure | 1,030 | 9.1 | 868 | 7.6 | - |  | - |  |
| Monozygotic | 2,993 | 26.5 | 3,469 | 30.3 | 152 | 69.1 | 457 | 49.0 |
| Same-sex dizygotic | 7,285 | 64.4 | 7,105 | 62.1 | 68 | 30.9 | 476 | 51.0 |
| Health status |  |  |  |  |  |  |  |  |
| Illnesses ^a^ | 8,704 |  | 9,409 |  | 205 |  | 850 |  |
| No | 8,094 | 93.0 | 8,924 | 94.8 | 203 | 99.0 | 809 | 95.2 |
| Yes | 610 | 7.0 | 485 | 5.2 | 2 | 1.0 | 41 | 4.8 |
| **Leisure-time physical activity** |  |  |  |  |  |  |  |  |
| Metabolic equivalent (MET) index |  |  |  |  |  |  |  |  |
| in 1975 | 10,328 | 2.7 (3.6) | 10,656 | 2.1 (2.5) | 216 | 2.8 (3.6) | 900 | 2.0 (2.1) |
| in 1981 | 9,788 | 2.9 (3.7) | 10,377 | 2.3 (2.5) | 210 | 2.6 (2.9) | 888 | 2.3 (2.2) |
| in 1990 | 5,621 | 3.2 (3.6) | 6,691 | 3.3 (3.2) | 173 | 3.4 (3.2) | 668 | 3.0 (2.6) |
| **Health-related factors in 1981** |  |  |  |  |  |  |  |  |
| BMI in 1981 | 9,768 | 24.4 (3.0) | 10,306 | 22.7 (3.2) | 208 | 22.7 (3.4) | 889 | 23.3 (3.5) |
| Smoking in 1981 | 11,308 |  | 11,442 |  | 209 |  | 933 |  |
| Never | 3,043 | 31.5 | 5,889 | 57.8 | 93 | 44.5 | 610 | 69.6 |
| Occasional | 384 | 4.0 | 240 | 2.4 | 13 | 6.2 | 20 | 2.3 |
| Former | 2,487 | 25.7 | 1,662 | 16.3 | 52 | 24.9 | 130 | 14.8 |
| Light | 581 | 6.0 | 895 | 8.8 | 10 | 4.8 | 43 | 4.9 |
| Medium | 1,632 | 16.9 | 1,082 | 10.6 | 25 | 12.0 | 52 | 5.9 |
| Heavy | 1,538 | 15.9 | 417 | 4.1 | 16 | 7.7 | 21 | 2.4 |
| Alcohol use in 1981 ^b^ | 9,672 |  | 10,146 |  | 210 |  |  |  |
| Never | 462 | 4.8 | 1,306 | 12.9 | 10 | 4.8 | 157 | 17.8 |
| Former | 329 | 3.4 | 763 | 7.5 | 9 | 4.3 | 60 | 6.8 |
| Occasional | 182 | 1.9 | 863 | 8.5 | 2 | 1.0 | 75 | 8.5 |
| Low | 7,299 | 75.5 | 7,038 | 69.4 | 169 | 80.5 | 577 | 65.5 |
| Medium | 836 | 8.6 | 125 | 1.2 | 13 | 6.2 | 7 | 0.8 |
| High | 361 | 3.7 | 37 | 0.4 | 6 | 2.9 | 4 | 0.5 |
| Very high | 203 | 2.1 | 14 | 0.1 | 1 | 0.5 | 1 | 0.1 |
| **Outcomes** |  |  |  |  |  |  |  |  |
| Deaths (1991-2020) | 4,291 | 37.9 | 2,664 | 23.3 | 12 | 5.5 | 255 | 27.3 |
| Biological aging |  |  |  |  |  |  |  |  |
| Age at blood-draw | - |  | - |  | 220 | 65.0 (8.5) | 933 | 63.2 (9.2) |
| PC-based DNAm GrimAge |  |  |  |  | 220 | 74.7 (7.3) | 933 | 70.0 (7.1) |
| DunedinPACE | - |  | - |  | 220 | 1.02 (0.13) | 933 | 0.97 (0.11) |
| SD, standard deviation; DNAm, DNA methylation; PC, Principal component. | | | | |  |  |  |  |
| ^a^ Self-reported physician-diagnosed angina pectoris, myocardial infarction, or diabetes in 1975 or 1981. | | | | | | |  |  |
| ^b^ High and very high classes were combined for further analysis. | | | |  |  |  |  |  |

| Table S4. Model fit of the latent profile models with different number of classes (n = 22,750). | | | | | | | |
| --- | --- | --- | --- | --- | --- | --- | --- |
| Classes | AIC | BIC | ABIC | VLMR | LMR | Class sizes | AvePP |
| 1 | 275443 | 275491 | 275472 | - | - | - | - |
| 2 | 232782 | 232887 | 232845 | <0.001 | <0.001 | 76.8%, 23.2% | 0.95, 0.93 |
| 3 | 221285 | 221446 | 221382 | <0.001 | <0.001 | 54.6%, 29.9%, 15.4% | 0.92, 0.84, 0.95 |
| 4 | 217829 | 218046 | 217960 | 0.227 | 0.230 | 38.7%, 36.7%, 13.4%, 11.1% | 0.86, 0.81, 0.78, 0.93 |
| 5 | 215064 | 215337 | 215229 | <0.001 | <0.001 | 34.9%, 34.2%, 12.7%, 10.0%, 8.3% | 0.79, 0.85, 0.73, 0.93, 0.77 |
| 6 | 212645 | 212974 | 212844 | 0.003 | 0.004 | 38.7%, 33.0%, 11.4%, 8.4%, 6.0%, 2.4% | 0.86, 0.78, 0.92, 0.82, 0.80, 0.94 |
| AIC, Akaike’s information criterion; BIC, Bayesian information criterion; ABIC, sample size-adjusted Bayesian information criterion; VLMR, Vuong-Lo-Mendell-Rubin likelihood ratio test; LMR, Lo-Mendell-Rubin-adjusted likelihood ratio test; AvePP, average posterior probabilities for most likely latent class membership. | | | | | | | |

| Table S5. The characteristics in the latent classes with different long-term leisure-time physical activity patterns in the population weighted by classification probabilities (n = 22,750). | | | | | | | | |
| --- | --- | --- | --- | --- | --- | --- | --- | --- |
|  | C1 Sedentary (13.4%) | | C2 Moderately active (36.7%) | | C3 Active (38.7%) | | C4 Highly active (11.2%) | |
|  | n | % or Mean (SD) | n | % or Mean (SD) | n | % or Mean (SD) | n | % or Mean (SD) |
| Sex | 3,040 |  | 8,355 |  | 8,809 |  | 2,545 |  |
| Male | 1,620 | 53.3 | 3,930 | 47.0 | 4,168 | 47.3 | 1,590 | 62.5 |
| Female | 1,420 | 46.7 | 4,425 | 53.0 | 4,641 | 52.7 | 955 | 37.5 |
| Age in 1975 | 3,040 | 31.8 (9.4) | 8,355 | 30.2 (8.9) | 8,810 | 29.8 (8.8) | 2,545 | 27.9 (8.4) |
| Health status |  |  |  |  |  |  |  |  |
| Diseases ^a^ | 2,249 |  | 6,690 |  | 7,139 |  | 2,035 |  |
| No | 2,073 | 92.2 | 6,272 | 93.8 | 6,726 | 94.2 | 1,947 | 95.7 |
| Yes | 176 | 7.8 | 418 | 6.2 | 413 | 5.8 | 88 | 4.3 |
| Education in 1981 | 2,980 | 7.5 (2.5) | 8,165 | 8.4 (3.1) | 8,593 | 8.8 (3.3) | 2,466 | 9.1 (3.4) |
| BMI in 1981 | 2,549 | 24.3 (3.7) | 7,402 | 23.6 (3.4) | 7,856 | 23.3 (3.3) | 2,266 | 23.1 (2.9) |
| Smoking in 1981 | 2,507 |  | 7,319 |  | 7,787 |  | 2,235 |  |
| Never | 1,069 | 42.6 | 3,208 | 43.8 | 3,580 | 46 | 1,075 | 48.1 |
| Occasional | 65 | 2.6 | 230 | 3.1 | 247 | 3.2 | 82 | 3.7 |
| Former | 457 | 18.2 | 1,515 | 20.7 | 1,683 | 21.6 | 493 | 22.1 |
| Light | 162 | 6.5 | 569 | 7.8 | 593 | 7.6 | 151 | 6.8 |
| Medium | 382 | 15.2 | 1,051 | 14.4 | 1,017 | 13.1 | 264 | 11.8 |
| Heavy | 372 | 14.8 | 746 | 10.2 | 667 | 8.6 | 170 | 7.6 |
| Alcohol use in 1981 | 2,500 |  | 7,325 |  | 7,757 |  | 2,237 |  |
| Never | 323 | 12.9 | 643 | 8.8 | 632 | 8.1 | 170 | 7.6 |
| Former | 136 | 5.4 | 381 | 5.2 | 457 | 5.9 | 119 | 5.3 |
| Occasional | 156 | 6.2 | 394 | 6.4 | 388 | 5.0 | 107 | 4.8 |
| Low | 1,657 | 66.3 | 5,316 | 72.6 | 5,698 | 73.5 | 1,666 | 74.5 |
| Medium | 130 | 5.2 | 350 | 4.8 | 364 | 4.7 | 117 | 5.2 |
| High | 51 | 2.0 | 163 | 2.2 | 141 | 1.8 | 43 | 1.9 |
| Very high | 47 | 1.9 | 78 | 1.1 | 77 | 1.0 | 15 | 0.7 |
| BMI, body mass index; SD, standard deviation. | | | | | | | | |
| ^a^ Self-reported physician-diagnosed angina pectoris, myocardial infarction, or diabetes in 1975 or 1981. | | | | | | |  |  |

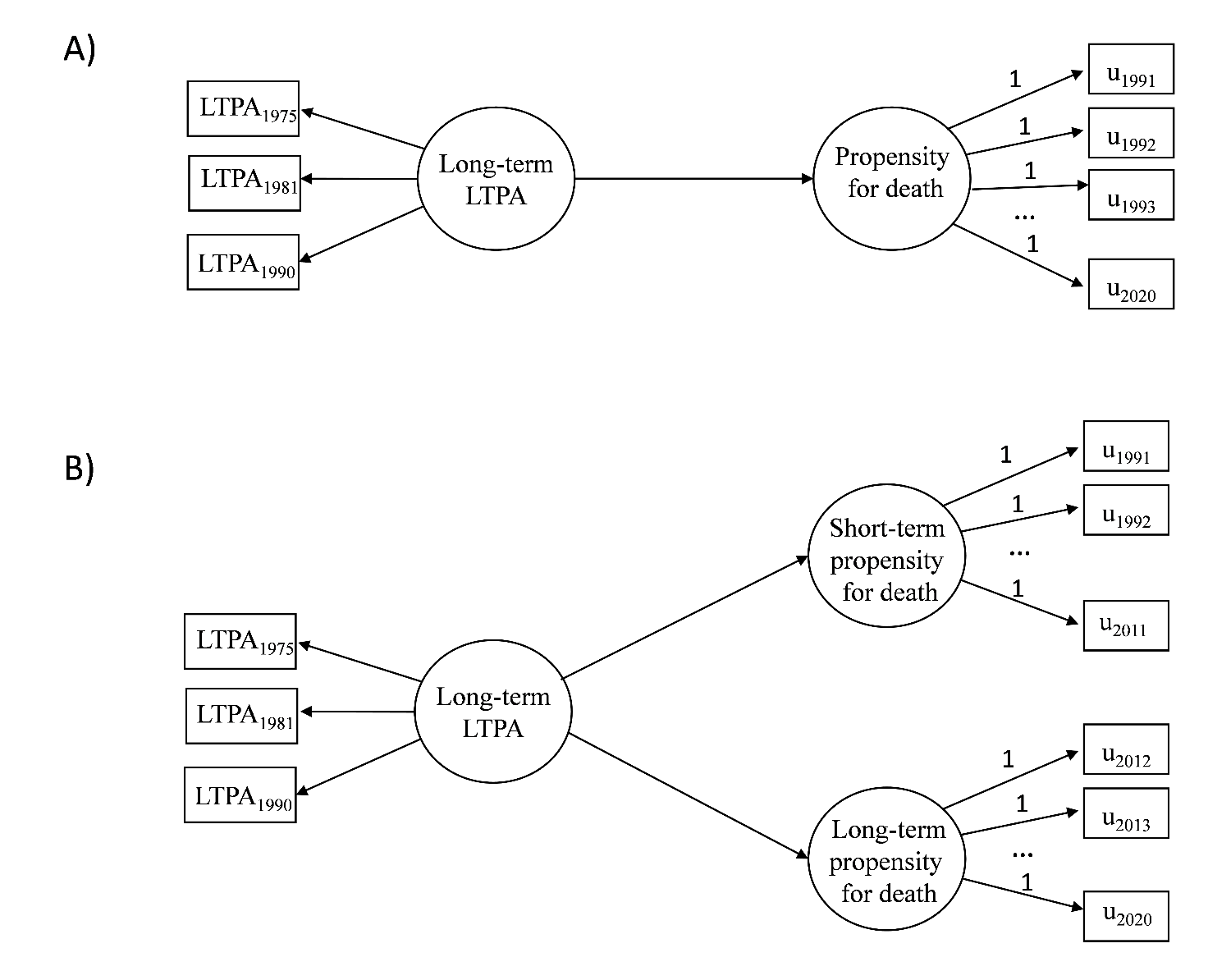

Figure S1. Path diagram of the discrete-time survival models for A) total mortality and B) short- and long-term mortality. Follow-up time was treated as time scale in the analysis. Circles denote latent variables and rectangles observed variables.

LTPA, leisure-time physical activity; u, discrete-time survival indicators.

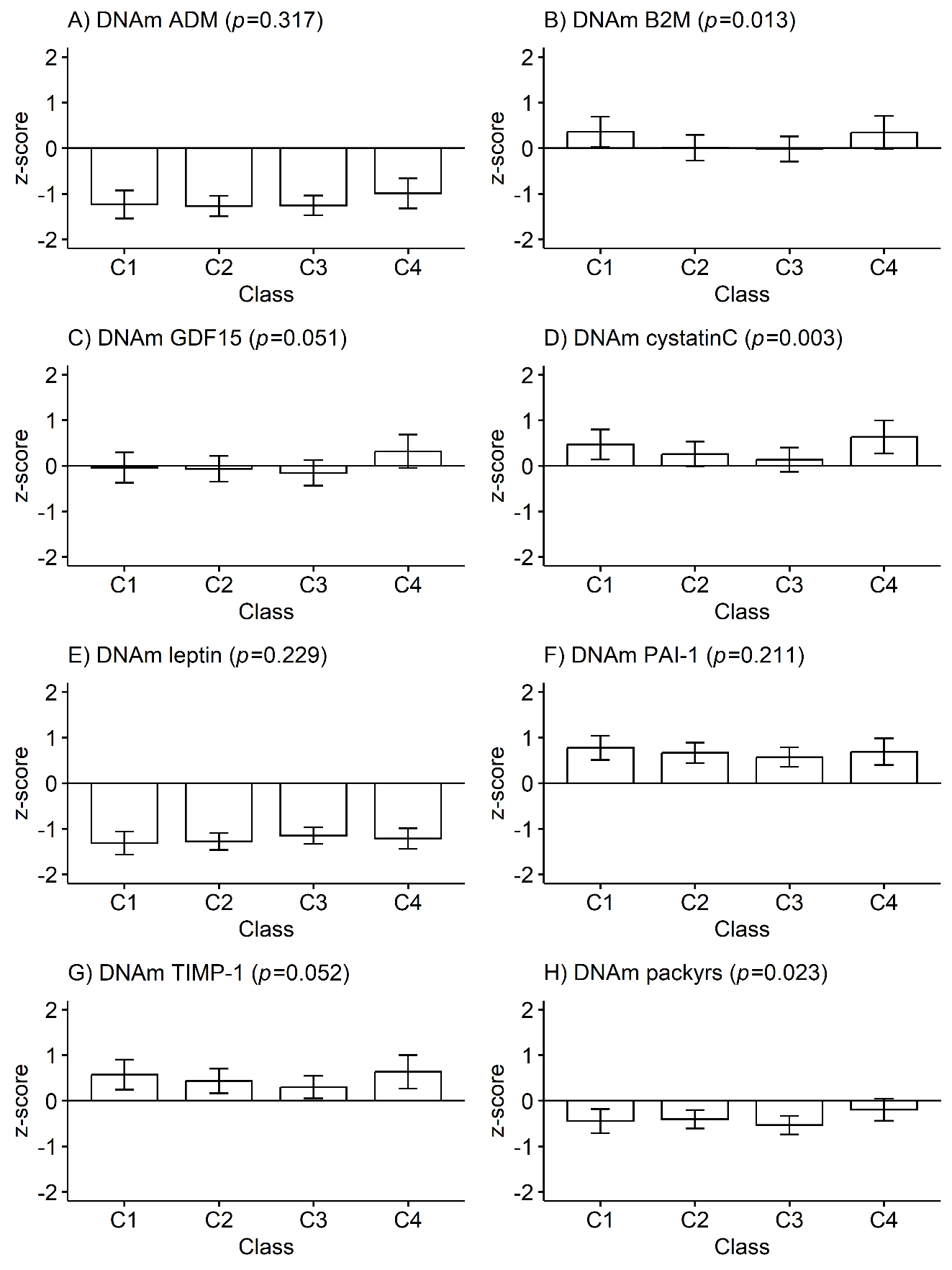

Figure S2. Mean differences in PC-based DNA methylation (DNAm)-based plasma proteins and smoking pack-years between the classes of long-term leisure-time physical activity (n = 1,153): A) DNAm adrenomedullin (ADM), B) DNAm beta-2 microglobulin (B2M), C) DNAm growth differentiation factor (GDF15), D) DNAm cystatin C, E) DNAm leptin, F) DNAm plasminogen activation inhibitor 1 (PAI-1), DNAm tissue inhibitor metalloproteinase 1 (TIMP-1), and H) DNAm smoking pack-years (packyrs). Means and 95% confidence intervals are presented. C1, Sedentary; C2, Moderately active; C3, Active; C4 Highly active. The model was adjusted for sex (female), age, health status, education years, body mass index, smoking, and alcohol use. P value from overall Wald test.

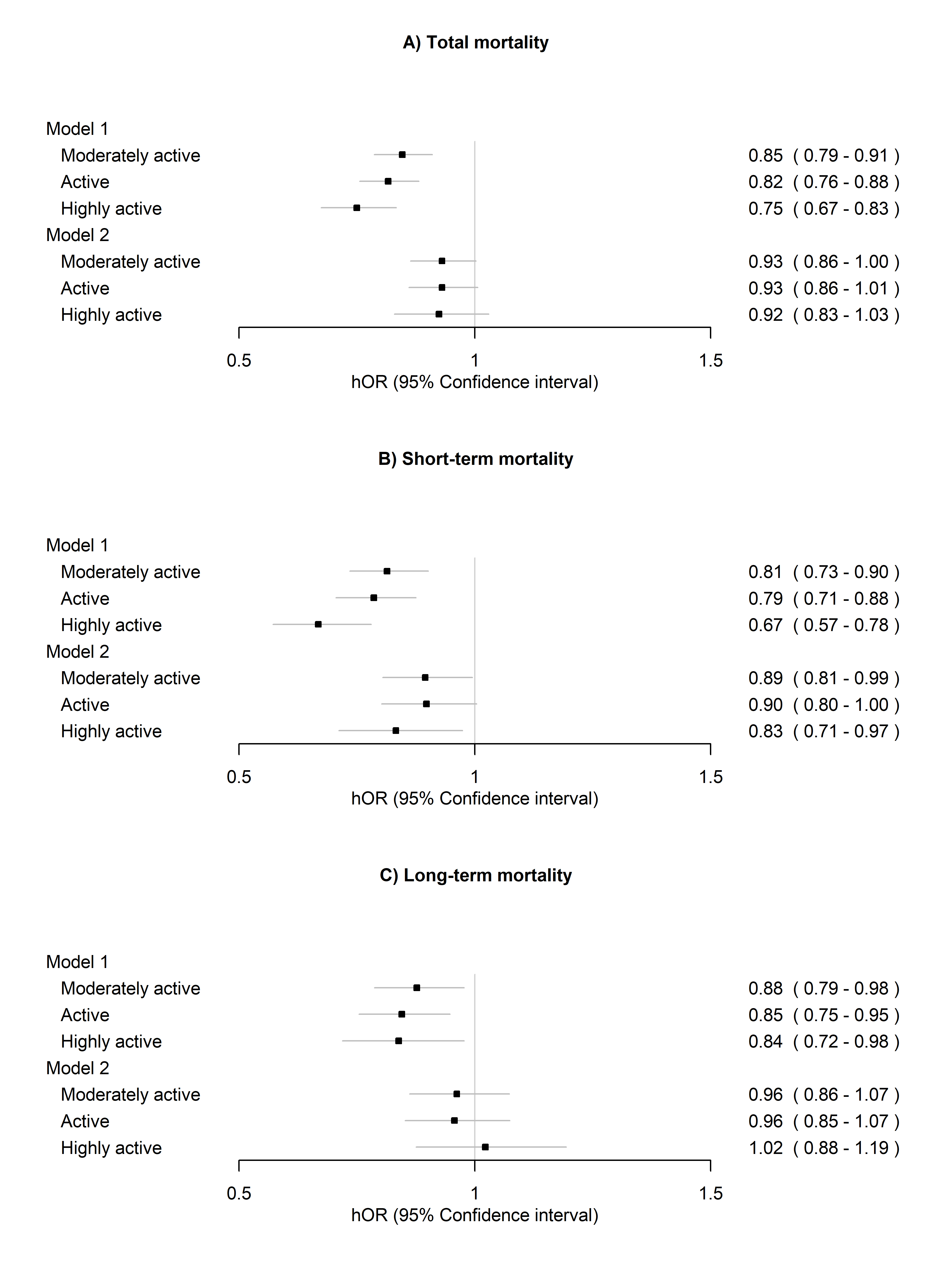

Figure S3. Associations of long-term leisure-time physical activity with A) total mortality, B) short-term mortality (1990–2011), and C) long-term mortality (2012–2020). Twins who did not report selected diseases were included in the analysis (n=17,018). Sedentary class was treated as reference.

Model 1 was adjusted for sex (female), age and health status.

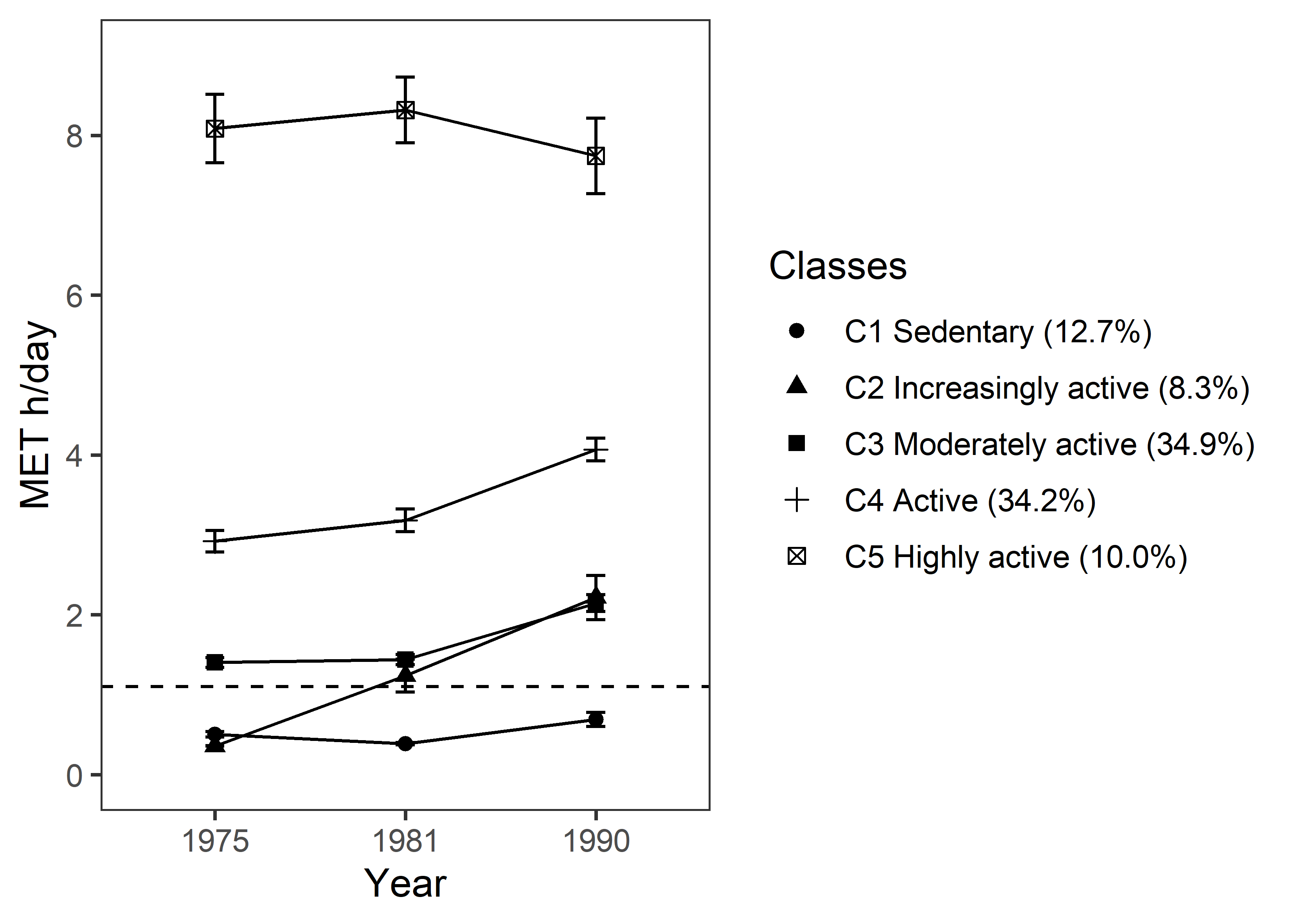

Figure S4. Sensitivity analysis: Latent profile solution with five classes (n = 22,750). Means of metabolic equivalent (MET) hours (h)/day and 95% confidence intervals are presented. The dashed line denotes World Health Organization guidelines for the recommended minimum amount of physical activity for adults (150 min of moderate intensity physical activity per week ~ 1.1 MET h/day).

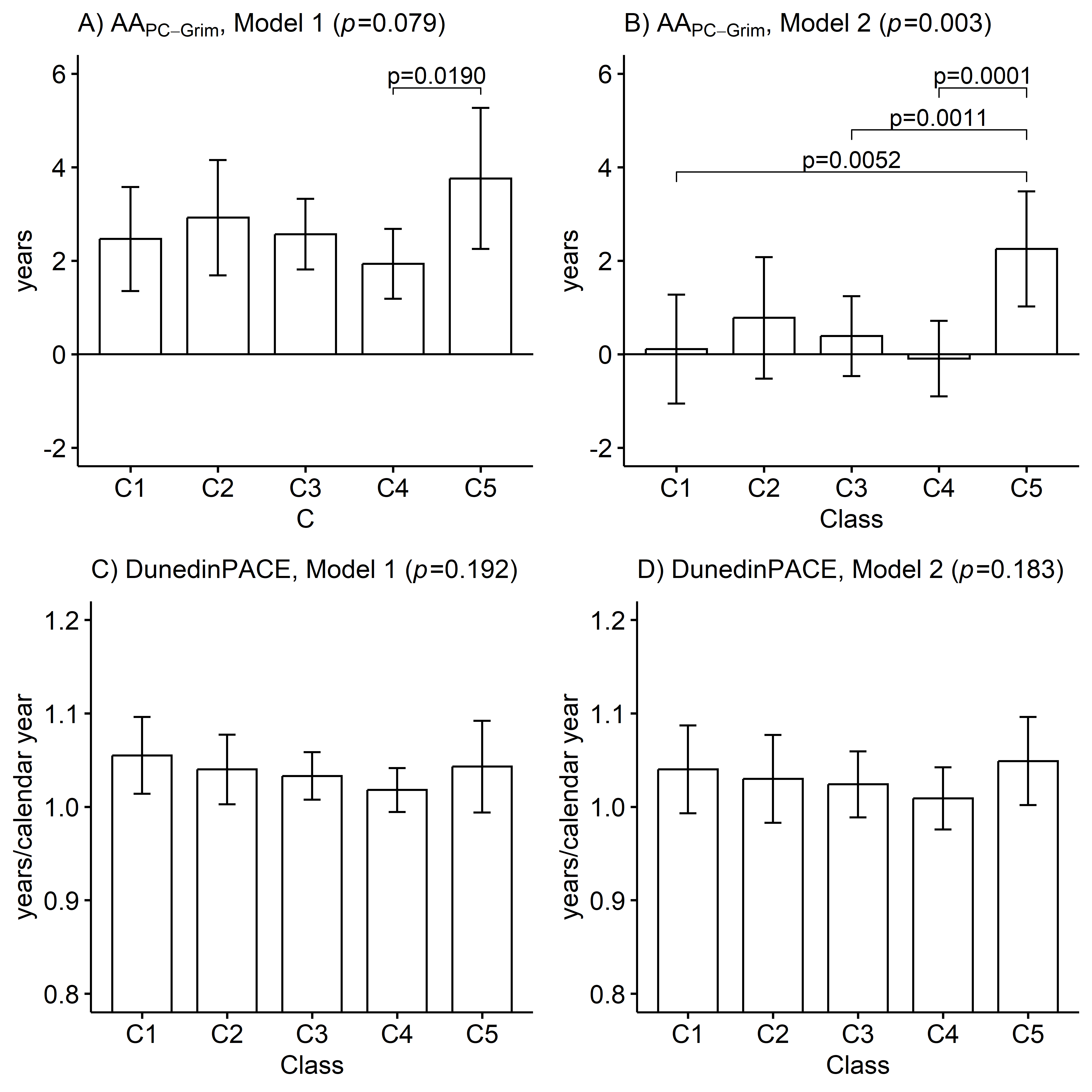

Figure S5. Sensitivity analysis: latent profile solution with five classes; mean differences between the classes of long-term leisure-time physical activity in biological aging measured with A–B) PC-based GrimAge and C–D) DunedinPACE (n=1,153). Means and 95% confidence intervals are presented. Model 1 was adjusted for sex (female), age, and health status, and Model 2 additionally for education, body mass index, smoking, and alcohol use. C1, Sedentary (8.0%); C2, Increasingly active (from sedentary to moderate) (7.7%); C3, Moderately active (39.3%); C4, Active (39.4%); C5 Highly active (5.6%); AA, Age acceleration. p value from Wald test.

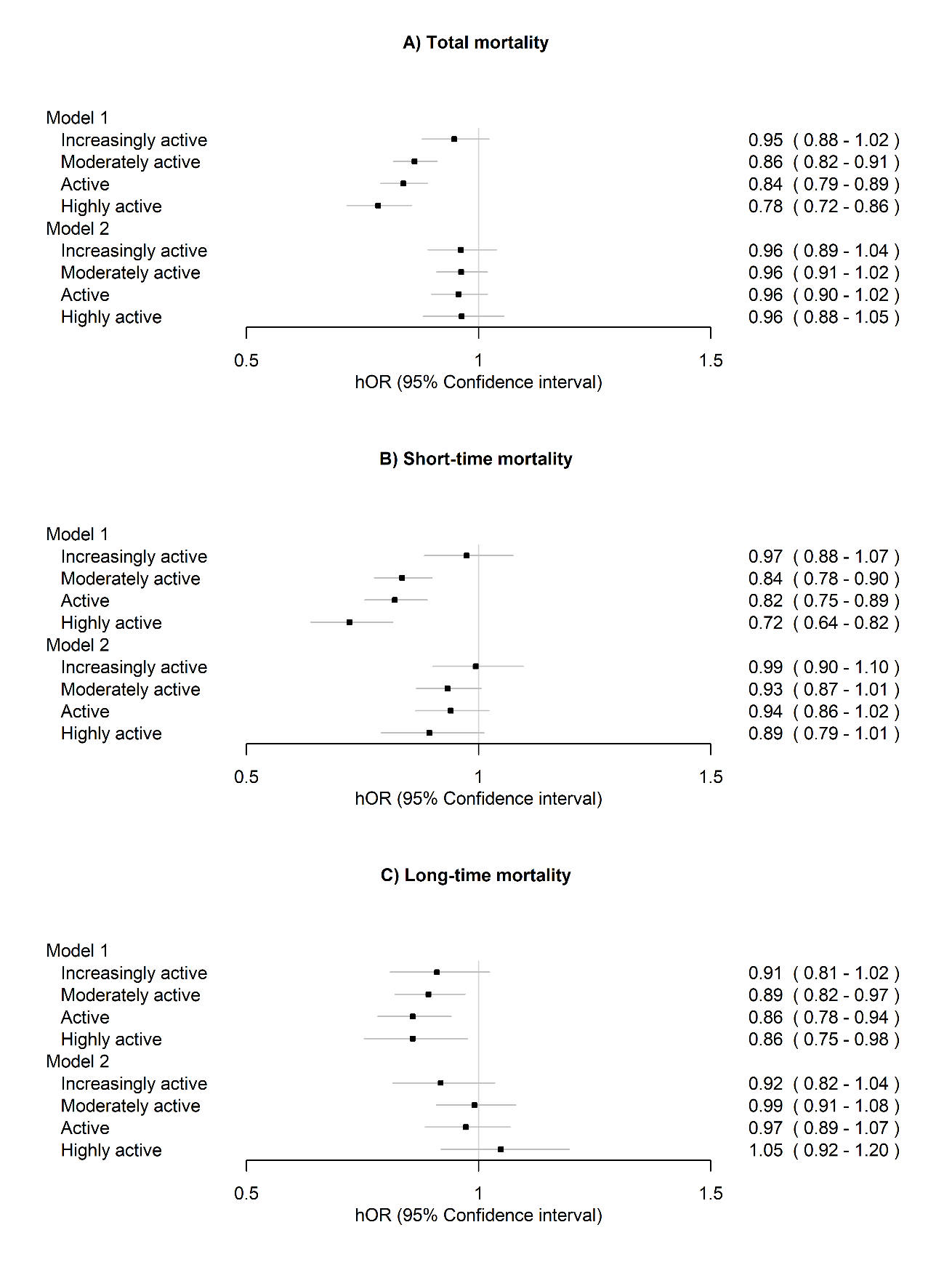

Figure S6. Sensitivity anaysis: Latent profile solution with five classes, the associations of long-term leisure-time physical activity with A) total mortality, B) short-term mortality (1990–2011), and C) long-term mortality (2012–2020) (n=22,750). Sedentary class was treated as reference.

Model 1 was adjusted for sex (female), age and health status.

Model 2 was additionally adjusted for education, body mass index, smoking, and alcohol use.

hOR, hazard odds ratio.

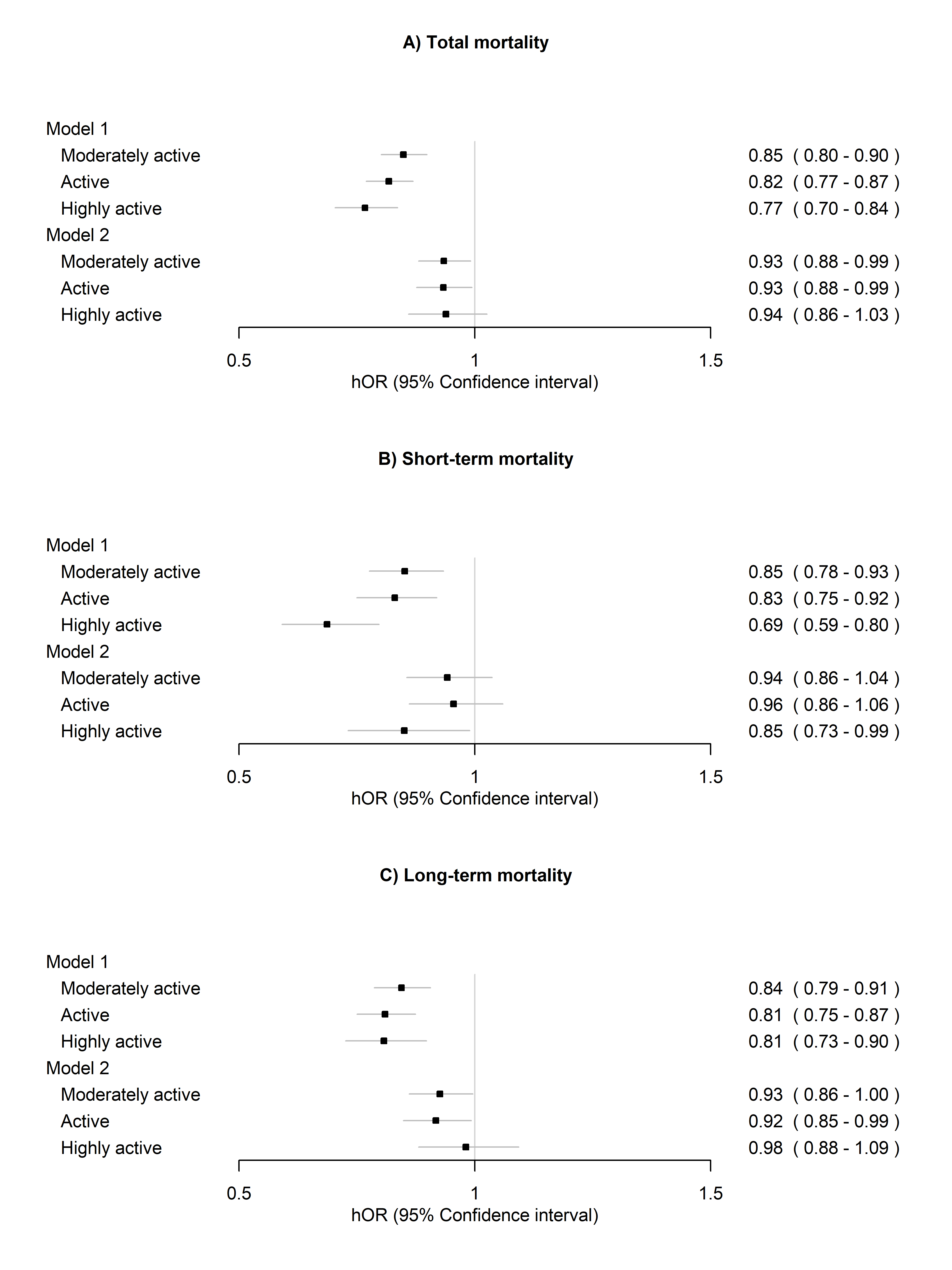

Figure S7. Sensitivity analysis: The associations of long-term leisure-time physical activity with A) total mortality, B) short-term mortality (1990–2006), and B) long-term mortality (2007–2020) (n=22,750).

Model 1 was adjusted for sex (female), age and health status.

Model 2 was additionally adjusted for education, body mass index, smoking, and alcohol use.
